# Poor knowledge of COVID-19 and unfavourable perception of the response to the pandemic by healthcare workers at the Bafoussam Regional Hospital (West Region - Cameroon)

**DOI:** 10.1101/2020.08.20.20178970

**Authors:** Jovanny Tsuala Fouogue, Michel Noubom, Bruno Kenfack, Norbert Tanke Dongmo, Maxime Tabeu, Linda Megozeu, Jean Marie Alima, Yannick Fogoum Fogang, Landry Charles A Nyam Rim, Florent Ymele Fouelifack, Jeanne Hortence Fouedjio, Pamela Leonie Fouogue Nzogning Manebou, Clotaire Damien Bibou Ze, Brice Foubi Kouam, Lauriane Nomene Fomete, Pierre Marie Tebeu, Jean Dupont Ngowa Kemfang, Pascal Foumane, Zacharie Sando, George Enownchong Enow Orock

## Abstract

**Background:** The World Health Organization has warned against a dramatic impact of COVID-19 in sub-Saharan Africa unless adequate response strategies are implemented. Whatever the strategy, the role of health care workers is pivotal. We undertook this study to assess knowledge of COVID-19 and perception of the response to the pandemic among the staff of a regional hospital in charge of COVID-19 patients in West Cameroon.

**Methods:** We used a convenience non probabilistic sampling method to carry out a survey with a self-administered questionnaire from April 14, 2020 to April 29, 2020 at the Bafoussam Regional Hospital (BRH). All the staff was invited to participate. Statistical analyses were done using Microsoft Excel 2010 and Epi-Info version 7.1.5.2 software.

**Results:** Response rate was 76.1% (464/610). Mean age (SD) and average work experience (SD) were 35.0 (8.9) and 8.4 (7.4) years respectively. Sex ratio (M/F) was 101/356. Nursing and midwifery staff (56.8%) and in-patients units (49.94%) were predominant. Knowledge on origin and transmission of SARS-CoV-2 was poor but knowledge of clinical signs and the role of laboratory tests were good. 53.2% of respondents said all therapeutic regimens are only supportive and only a third of them trusted drugs recommended by health authorities. For 36.9% of respondents, herbal remedies can prevent/cure COVID-19. 70% of staffs felt they were not knowledgeable enough to handle COVID-19 cases. 85.6% of respondents thought the BRH had insufficient resources to adequately respond to COVID-19 and 55.6% were dissatisfied with its response to the pandemic (weaknesses: medicines/technologies (74.5%), service delivery (28.1%), human resource (10.9%)). 68% of staff felt insufficiently protected on duty and 76.5% reported that the pandemic significantly reduced non-COVID-19 services. 85.5% said they complied with preventive measures while in the community. For 44% of respondents Cameroonian regulations on COVID-19 corpses should be made more culture-sensitive. 51.2% of respondents were against vaccine trial in their community.

**Conclusion:** Knowledge of COVID-19 was poor and perception of the response to the pandemic was unfavorable.

## Introduction

SARS-CoV-2 and resulting coronavirus disease –2019 (COVID-19) originating from China (first case reported on December 8, 2019) was declared a global pandemic by the World Health Organization (WHO) on March 11, 2020 [1, 2]. The dramatic and rapid multidimensional (sanitary, social, economic, environment and political) impacts of COVID-19 worldwide has led several public health experts to rate it as the worst public health emergency since the Spanish flu in 1918 [3]. Considering the unprecedented and unforeseen rapid overwhelming of robust health systems and resilient economies of high-income countries by the pandemic and the structural weaknesses of African health systems, scientific and regulating organizations (including WHO) predicted a devastating COVID-19 wave in (Sub-Saharan) Africa (SSA) if the magnitude was to be similar [4, 5]. Fortuitously, the pandemic in Africa occurred several weeks behind Europe and Asia and has a slower transmission [5, 6]. (Sub-Saharan) African countries have therefore had more time to shape their containment and mitigation strategies under the guidance of the Africa Center for Disease Controls and assistance of the WHO [2, 3, 5, 7]. In Cameroon the first COVID-19 case was reported in the capital city (Yaounde) on March 6, 2020 and despite measures put in place by the government as from March 18, several cases were already reported in Regions [8]. Like several African countries the Cameroonian health system preparedness was not up to the magnitude of the pandemic [2]. The first COVID-19 case recorded in the West Region was admitted at the Bafoussam Regional Hospital (BRH) on March 18, 2020 and took (local) health authorities by surprise [9]. This resulted in a catch-up response rather than the expected anticipative response [8]. This is further illustrated by the quarantine imposed to the whole staff of a primary health care facility in Bafoussam (alongside its temporary closure) following a six-day hospitalization of COVID-19 symptomatic patient in an ordinary ward [10]. Whatever the strategy adopted by governments against COVID-19 frontline health care workers (HCWs) play a critical role and deserve special attention [6, 11 –15].

The goals of this study were to (i) assess the knowledge of HCWs regarding COVID-19 and (ii) determine their perception of the response to the pandemic at a regional level in (West) Cameroon.

## Materials and Methods

### Study setting

The Bafoussam Regional Hospital (BRH) is a tertiary 200 bed-capacity hospital, of 5 level health care system, that serves as the top reference health care facility for the West Region of Cameroon (roughly 2,250,000 inhabitants according to official projections [17]). It has been chosen by authorities to serve as the lone center for management of symptomatic COVID-19 cases in the Region.

### Study design and participants

A cross-sectional analytical survey was carried out among the staff of the BRH from April 14, 2020 to April 27, 2020. That was approximately one month after the first symptomatic COVID-19 patient recorded in the Region was admitted at the BRH [8, 9]. Prior to the survey, administrative approval and ethical clearance were obtained from hospital management and ethical board respectively. The sample was non-probabilistic and convenient. A pre-tested and self-administered paper questionnaire was distributed to all staff members via routine administrative channels. They were asked to fill and return the questionnaire as soon as possible. Several electronic reminders were sent to participants.

### Questionnaire

The self-designed questionnaire was validated by 6 reviewers (biologists, public health specialists, obstetricians-gynecologists, infectious disease specialists, intensivists, epidemiologists, pathologists) and a pre-test was carried out on 15 participants from Cameroon. Responses to the pre-test were not included in data analysis but were used to improve the questionnaire. Data were grouped into three categories: (i) socio-demographical and professional characteristics (age, sex, qualification, position, department, years of work experience, marital status, size of household, religion), (ii) knowledge of COVID-19 (awareness of COVID19-like pandemics, origin and transmission of Severe Acute Respiratory Syndrome Coronavirus-2 (SARS-CoV-2), clinical signs and severity of COVID-19, role of laboratory diagnosis, mother-to-child transmission, susceptibility to overt disease, efficacy of certain drugs (chloroquine, azithromycine, herbal remedies), management of COVID-19 corpses, impact of climate on transmission of SARS-CoV2) and (iii) perception of the response to the pandemic (observance of hygiene, barriers and distancing measures(HBDM) in communities and inside the BRH, weaknesses and satisfaction with the BRH institutional response to COVID-19, impact of the pandemic on routine hospital activities and opinion on eventual local research for a vaccine).

### Statistical analysis

Data were entered using the Microsoft Excel 2010 software and then imported and analysed using Epi-Info 7.1.5.2 software to generate tables of frequencies or 2×2 tables. Khi2 was used when necessary to compare proportions or Student-test (t-test) when necessary to compare means (replaced by Wilcoxon-test when abnormal distribution). The graphs were also generated using Microsoft Excel 2010 software.

## Results

Of the 610 questionnaires distributed 467 were returned among which 3 were poorly filled and excluded. Demographical and professional characteristics of respondents are shown in Table 1. Median age (range) was 33.0 (20 – 70) years and nursing and midwifery staff made up 56.8% of respondents. 49.9% of respondents were affiliated to in-patients units. The median number of years of work experience was 7 (3 – 48) years.

**Table 1.** Demographical and professional characteristics of respondents.

| Characteristics |  |
| --- | --- |
| **Proportion of respondents n/N (%)** | **464/610 (76.1)** |
| **Sex Ratio M/F,** | 101/356 |
| **Mean Age (SD)** | 35.0(8.9) |
| **Median Age (Range)** | 33.0 (20-70) |
| **Qualifications n/N (%)** | **458 / 464 (98.7)** |
| Nurses, midwives, and assimilated workers n (%) | 260 (56.8) |
| Biologists, and assimilated workers n (%) | 83 (18.1) |
| Administrators, and assimilated workers n (%) | 54 (11.8) |
| Water, sanitation and hygiene (WASH) staff and assimilated workers n (%) | 31 (6.8) |
| Physicians, Pharmacists, and Dentists n (%) | 30 (6.5) |
| **Departments n/N (%)** | **451/464 (97.2)** |
| In-patient units n (%) | 225(49.9) |
| Reception and out-patient units n (%) | 140 (31.0) |
| Administrations n (%) | 53 (11.7) |
| WASH and Mortuary n (%) | 33 (7.3) |
| **Years of work experience n/N(%)** | **443/464 (95.5)** |
| Mean (SD) | 8.4 (7.4) |
| Median (Range) | 7.0 (3-48) |
| Mode | 1.0 |
| **Senior position n/N(%)** | 137/409(33.5) |
| **Member of COVID-19 task force n/N (%)** | 12/464(2.6) |
| **Married (or in couple) n/N (%)** | 289/464(62.3) |
| **Size of the household (N)** | **281** |
| Mean (SD) | 5.6 (3.4) |
| Median (Range) | 5.0(3.5-19) |
| ≤ 5 members n (%) | 152(54.1) |
| > 5 members n (%) | 129(45.9) |
| **Religion (N)** | **464** |
| Christianism n (%) | 429(92.5) |
| Islam n (%) | 15(3.2) |
| Animism n (%) | 15(3.2) |
| Others n (%) | 5(1.1) |

SD: Standard Deviation; M: male; F: female.

Respondents’ knowledge towards COVID-19 is described in Table 2. Only 15.9% of respondents correctly identified the origin of SARS-CoV-2 and 35.3% correctly identified transmission routes. Respondents recognised 7.6±2.4 COVID-19 symptoms. 36.9% of respondents said herbal remedies protects against COVID-19. For 33.1% of respondents Chloroquine/Hydroxychloroquine is ineffective to prevent/cure COVID-19. 88.2% of respondents said COVID-19 corpses are highly contagious and should be buried at shortest. For 87.5% of respondents, public authorities should be involved in burying COVID-19 corpses while 44.1% thought family members alone should bury the corpse.

**Table 2.**
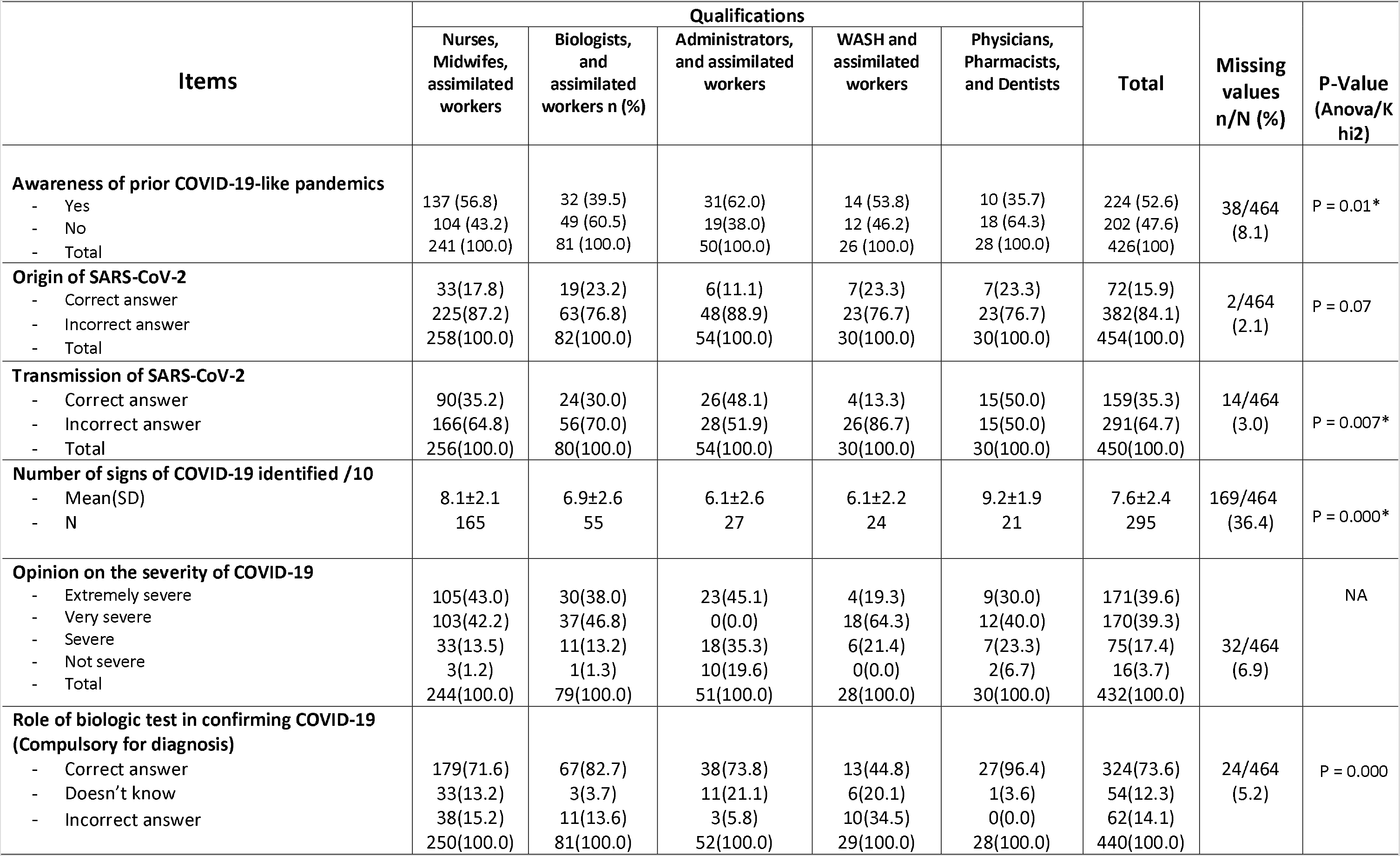
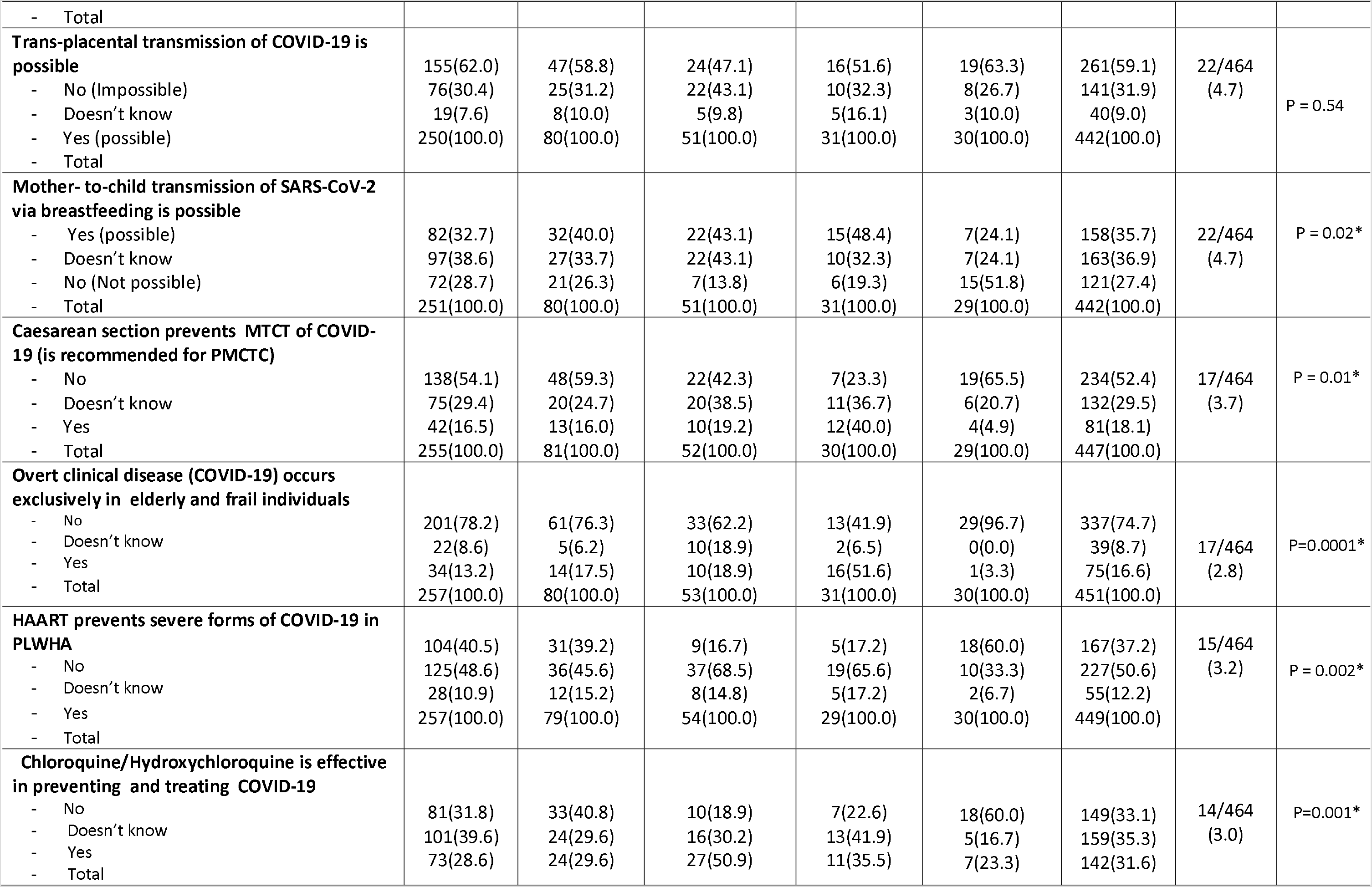
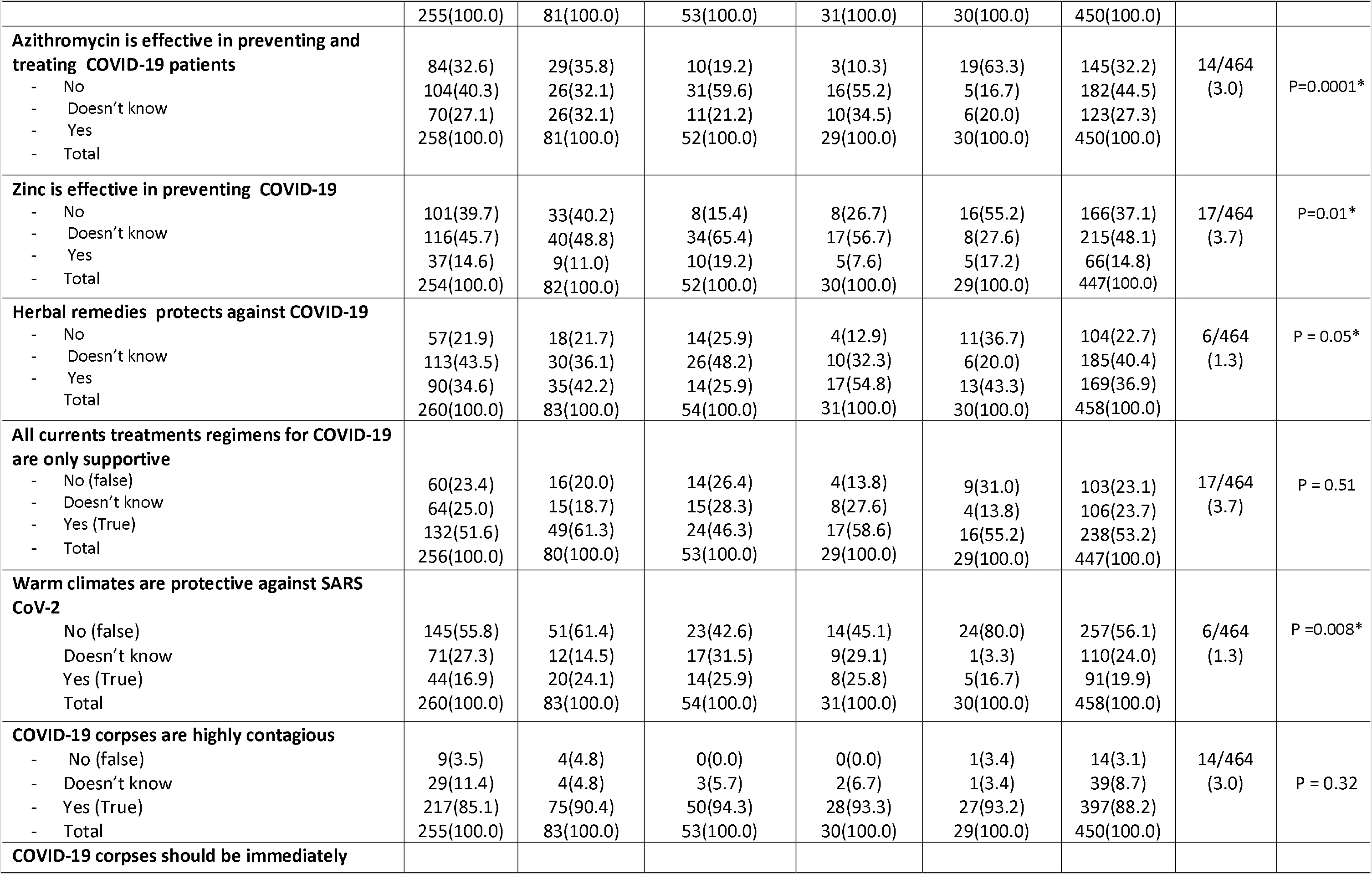
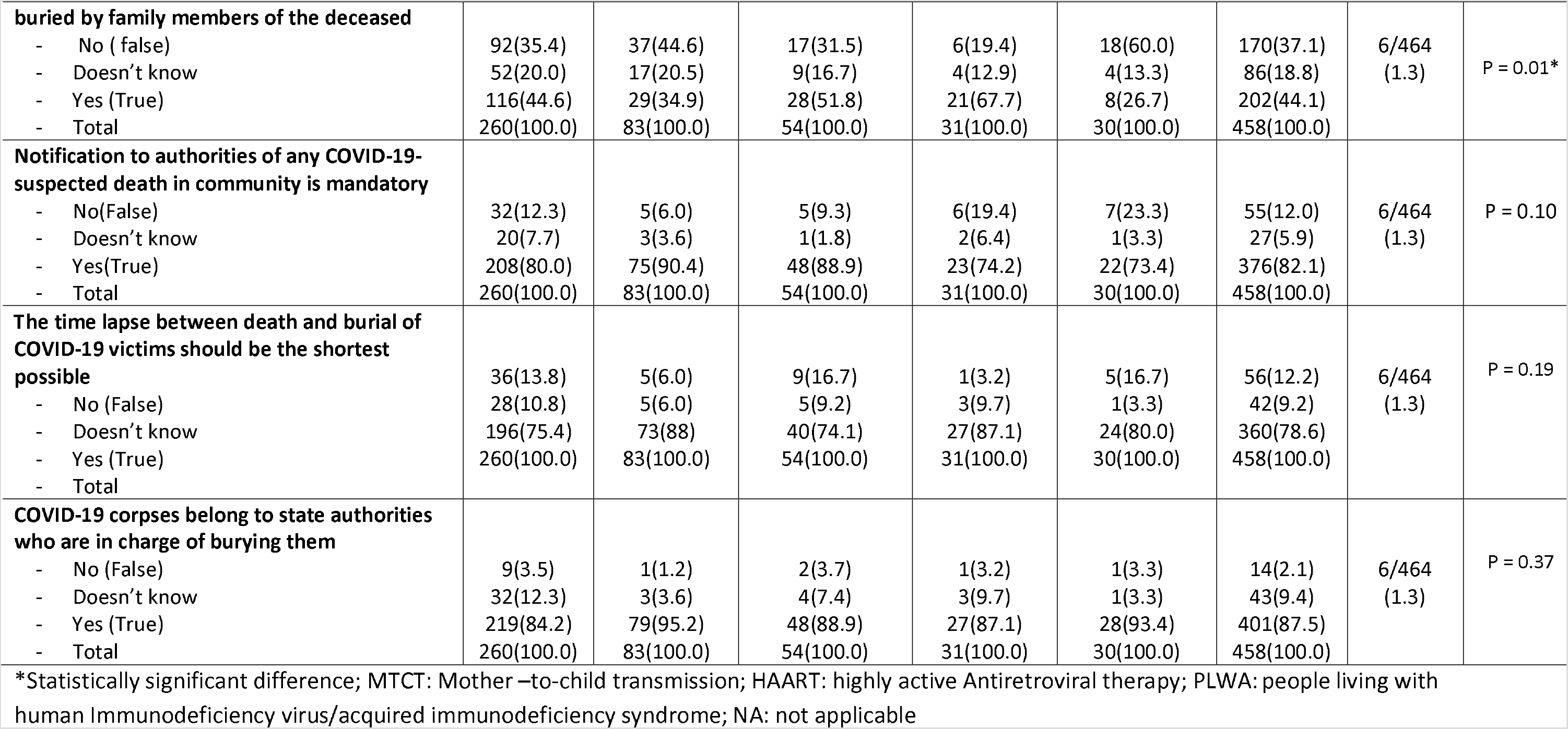
Knowledge of respondents towards COVID-19.

Table 3 summarizes of the response to COVID-19 by respondents. More than half (55.6%) of HCWs at the BRH were not satisfied with their hospital response to COVID-19 and 85.6% of HCWs thought its resources were insufficient to ensure an adequate response. As shown on Fig 1 perceived weaknesses of the BRH response to COVID-19 belonged to the following categories: medicines and technologies (74.5%), service delivery (28.1%) and human resource (10.9%). 70% of respondents said they were not Knowledgeable enough on how to handle COVID-19 cases and only 32% of them said they were not sufficiently protected (with personal protective equipment) in daily tasks. 51.2% of respondents were against vaccine trials in their community for the following reasons (Fig 2): vaccine trials represent a public health threat (33.7%); mistrust in state authorities to properly monitor the trials (33.2%) and conspiracy against Africans (22.4%).

**Table 3.**
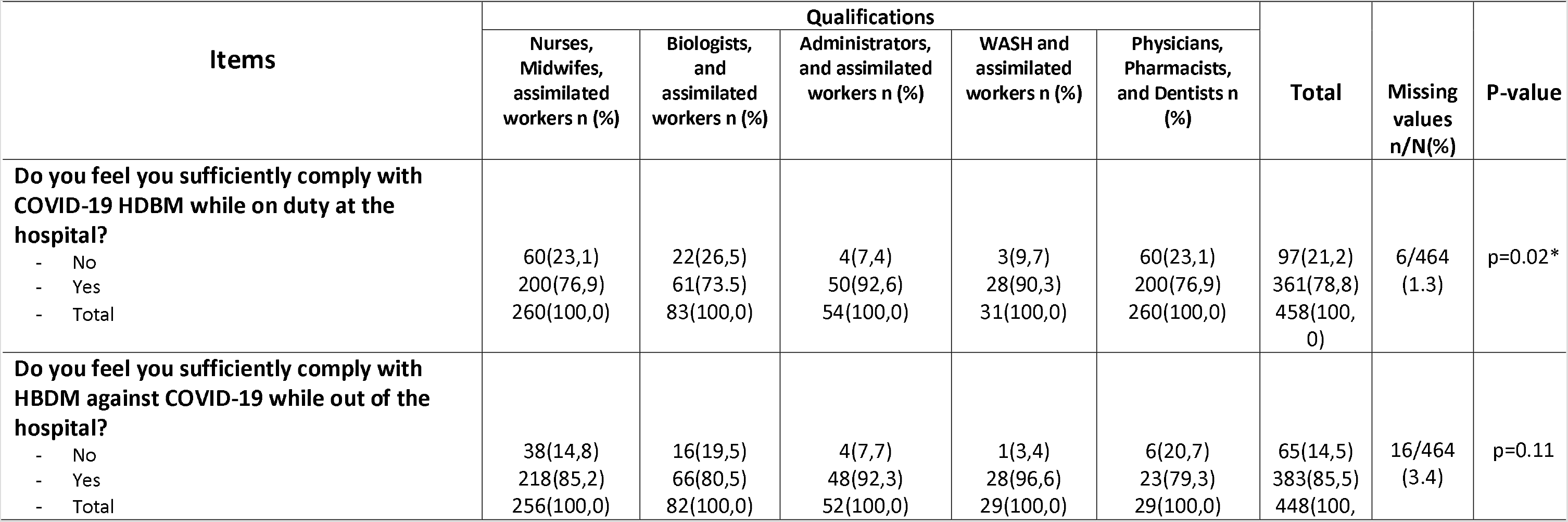
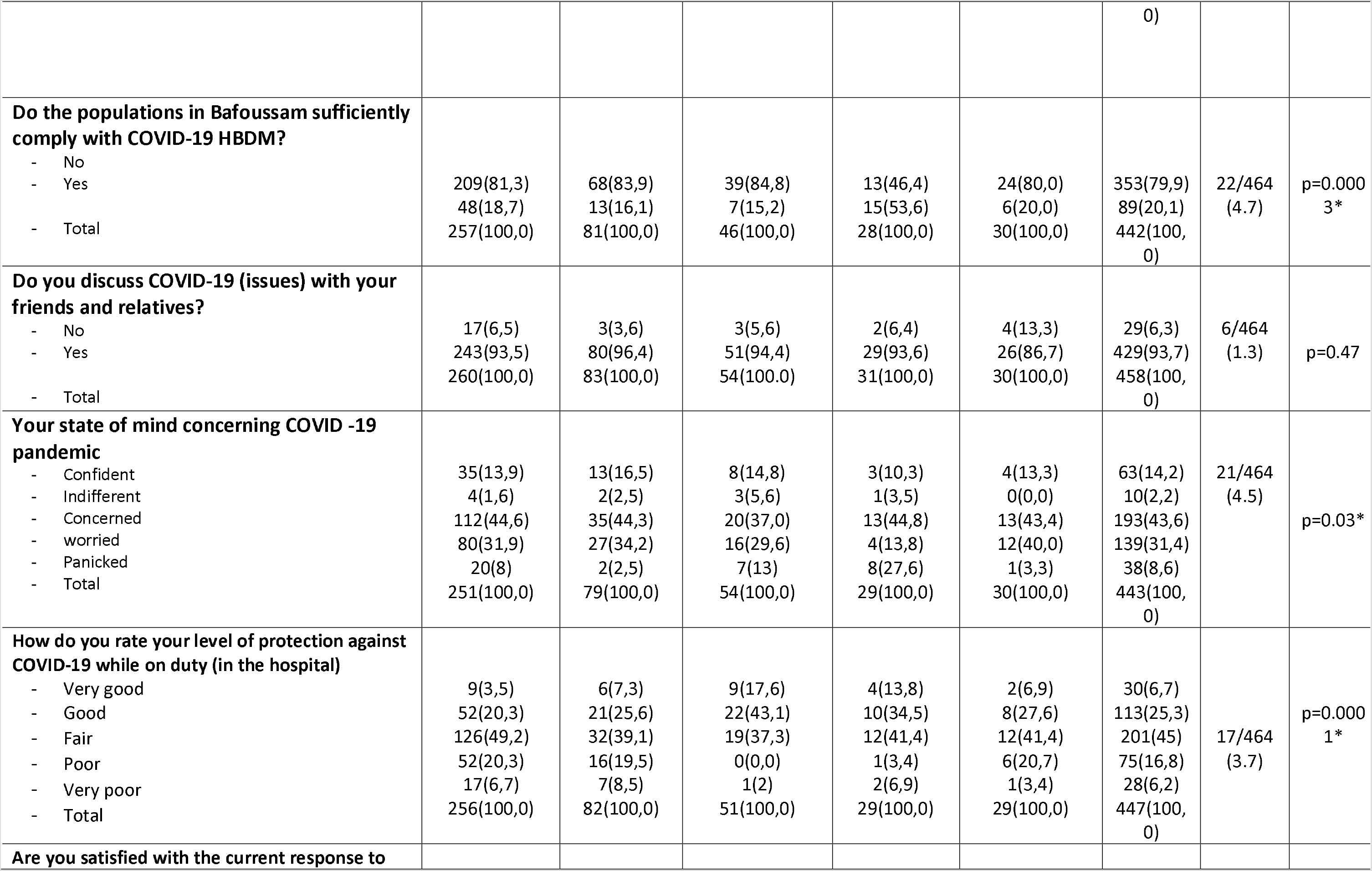
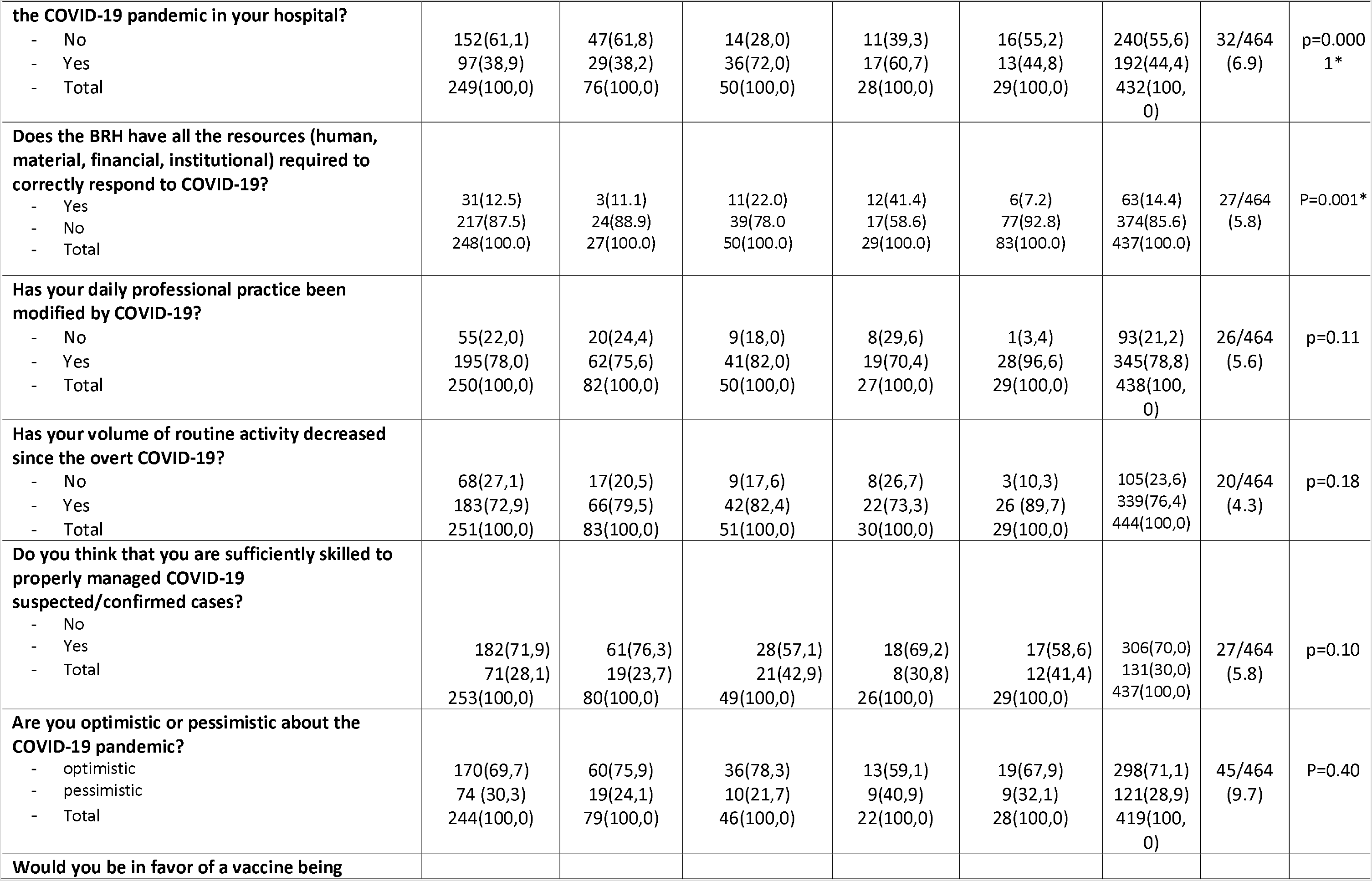
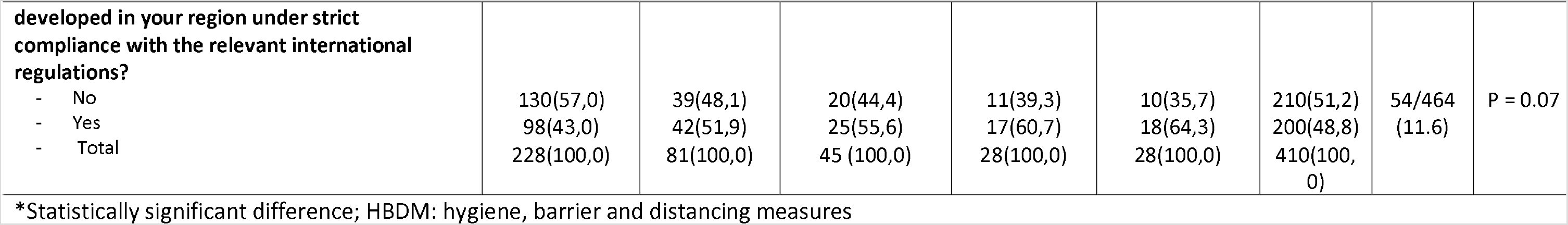
Perception of the response to COVID-19 by respondents.

**Figure 1.**
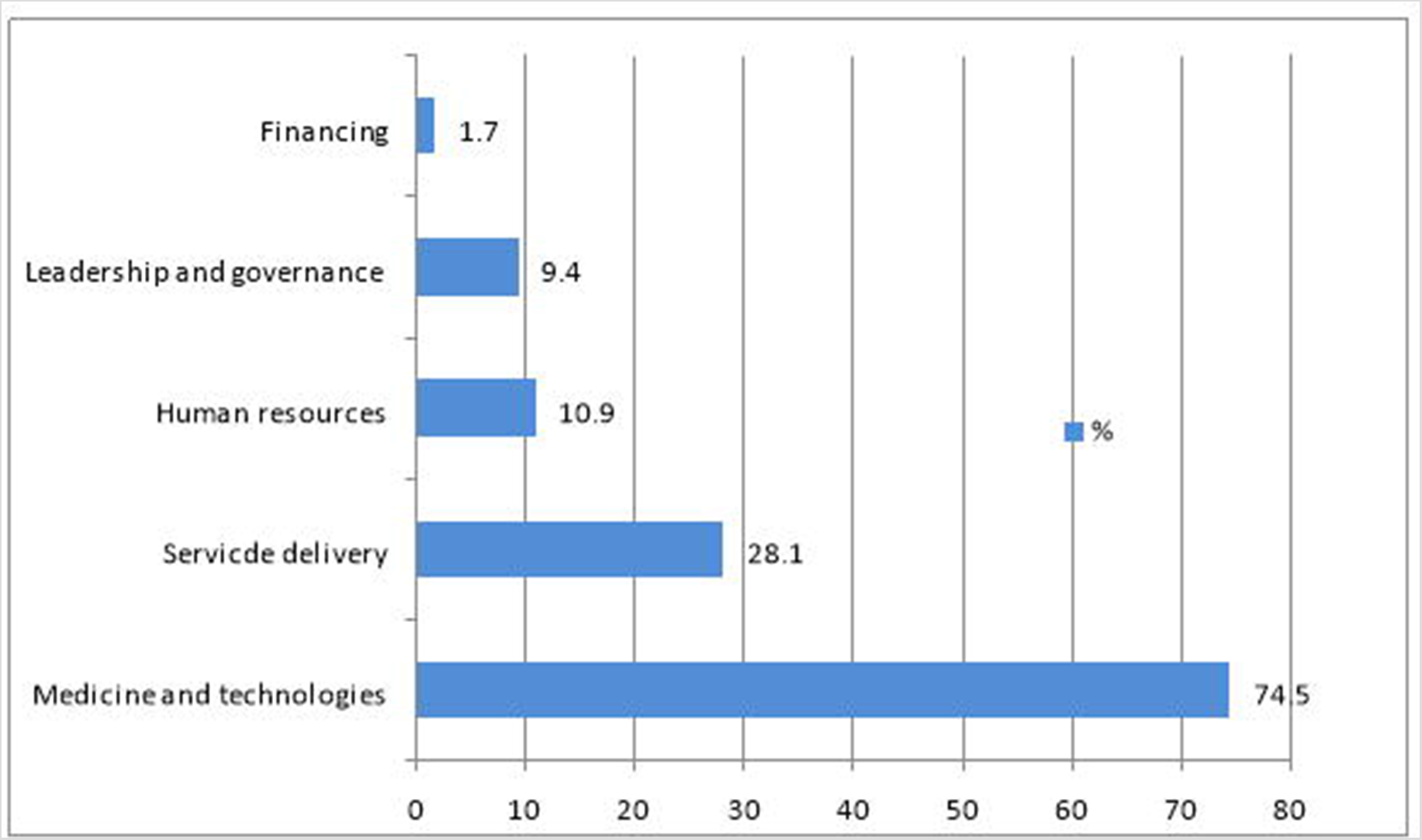
Perceived weaknesses of the Bafoussam Reeinal hospital response to COVID-19 by respondents

**Figure 2.**
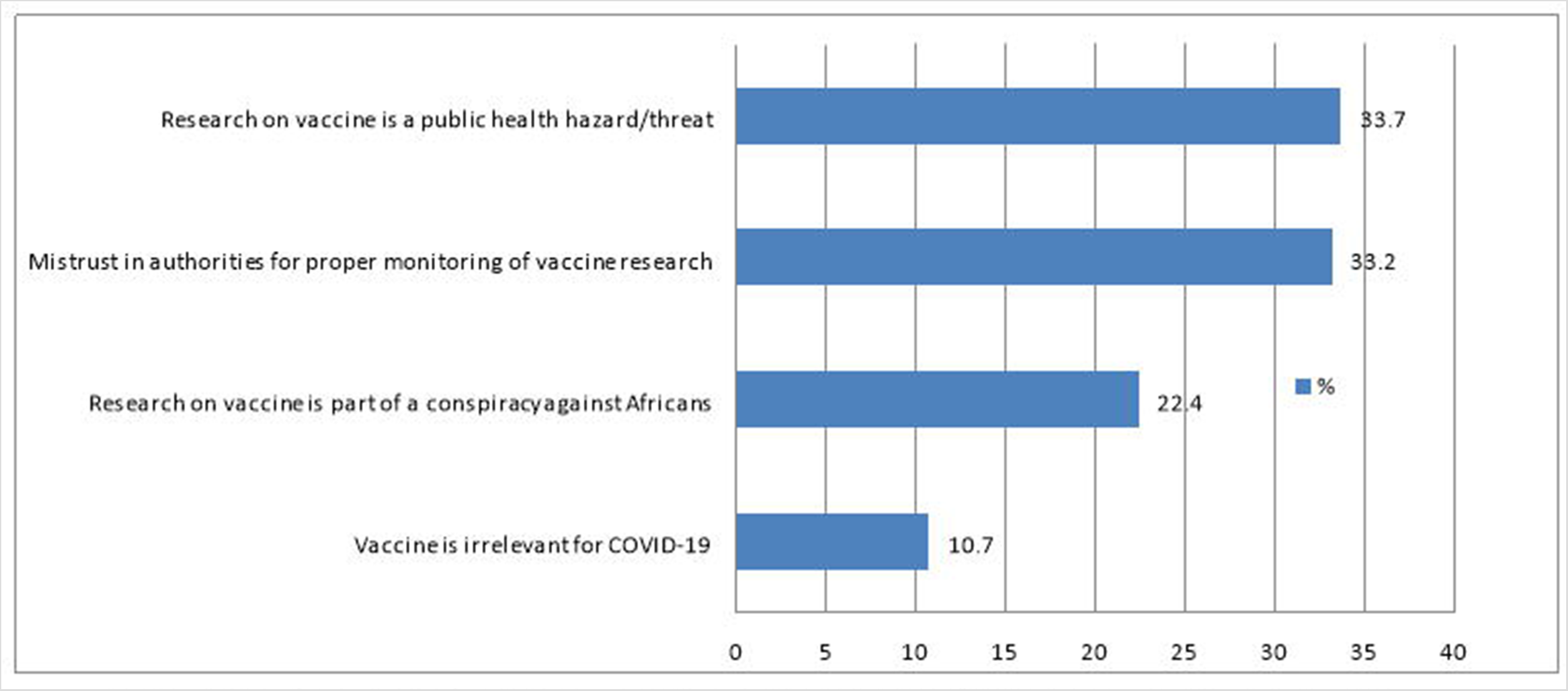
Motivations of opposition to local research for anti-COVID-19 vaccine among respondents

## Discussion

The response rate in this survey was satisfactory. The low M/F sex ratio observed (Table 1) is explained by the high proportion of nursing and midwifery staff which is characterised by a predominance of female workers in SSA [17]. Mean age of the respondents reflects the age distribution of health care workforce in the public sector in Cameroon [18]. The large sample size, the diversity of qualifications and departments ensures that results give near accurate trends in perception and knowledge of the pandemic in west Cameroon.

About half of respondents were aware that current COVID-19 pandemic is not the first in its kind, but very few of them (15.9%) correctly identified the origin of SARS-CoV-2 (Table 2); we found that only one third of respondents correctly identified transmission routes of SARS-CoV-2. Huang et al. reported poor knowledge among nursing staff in a Chinese hospital in charge of COVID-19 while Kamate et al. reported good knowledge of COVID-19 among dental practitioners worldwide with performance increasing with qualifications [19, 20]. Given the proximity of the oral cavity with contagious secretions of SARS-CoV2, it is plausible that dental practitioners have a higher risk perception that motivates accurate learning of transmission routes [20].

Flattening the curve of COVID-19 can’t be achieved unless HBDM are sufficiently observed in communities; this implies a good knowledge of transmission routes (the rationale for HBDM) that are better taught to (Sub Saharan) African populations by HCWs than governmental awareness actions especially where there is generalised mistrust and suspicion of corruption *vis-à-vis* public actions [1, 6]. The poor knowledge of transmission routes of SARS-CoV-2 by HCWs at the BRH should therefore be addressed for a more efficient communication on COVID-19 in the communities.

Up to one fifth of respondents wrongly thought that warm climate limits SARS-CoV-2 transmission with a significant difference between clinicians (nurses and physicians) and non-clinicians (administrators, biologists and WASH personnel) in favour of the previous. That belief has been one of the numerous arguments to explain the lower risk exposure and the lower transmission dynamics in SSA [4, 5, 21, 22]. Now that it is evident that climate doesn’t affect transmission (current surges in tropical countries during summer), we hope that the quarter of respondents with no opinion on the question will take position.

At the time of this survey there was no evidence of trans-placental transmission of SARS-CoV-2 (and prophylactic caesarean section was therefore not recommended) but breastfeeding was recommended by scientific bodies under strict observance of HBDM [23, 24]. Three fifths of respondents (without significant difference between qualifications categories) thought that trans-placental transmission was not possible while a third of them “didn’t know”. Only 18.1% of respondents would have recommended a prophylactic caesarean section while 52.4% were of the opinion that vaginal delivery doesn’t increase the risk of maternofoetal transmission of SARS-CoV-2. Clinicians (nurses and physicians) were significantly more likely to refrain from prophylactic caesarean section, an attitude in compliance with practice guidelines [23, 24]. A third of respondents would have (wrongly) advised against breastfeeding to prevent mother-to-child transmission of SARS-CoV-2 while another third had no answer to that question. There was a significant difference among categories, clinicians (nurses and physicians) being more in favour of breastfeeding. The proportions of clinicians (nurses and physicians) with no opinion on mother-to-child transmission of SARS-CoV-2 (ante-, per- or post-partum) were too high for an end-of-chain referral hospital. Indeed, without national clinical guidelines, dubious practitioners will intuitively tend to choose the most cautious options, namely (groundless) caesarean-section and formula-feeding despite their known risk in LMICs [25–28]. Regular knowledge updates of maternity staff on this emerging disease could be a solution.

Sorting of patients at the entrance of health facilities is paramount to avoid massive contamination of staff and other patients. An effective triage is based on good knowledge of clinical signs of COVID-19. We found that HCWs of the BRH in direct contact with patients (nurses and physicians) had a good knowledge of clinical signs of COVID-19. This combined with the fact that almost all (96.3%) of them acknowledged the severity of COVID-19 is a first step towards strict observance of HBDM on duty in a context marked by shortages of personal protective equipment (PPE) and absence of rapid diagnostic tests (at the time of this survey) [6, 7, 29, 30].

A majority of respondents (83.4%) knew that overt COVID-19 was not exclusively seen in elderly and frail individuals. This is reassuring and prevents social isolation of elder people out of stigma considering their very important social roles in SSA [31].

National guidelines for management of COVID-19 suspect cases were to isolate and treat them while waiting for confirmation with biologic test. At the time of the survey only Real time Polymerase Chain Reaction (RT-PCR) was available in a single laboratory throughout Cameroon to identify SARS-CoV-2. Results took several days to be available in regional hospitals [30]. Meanwhile, radiologic exams were much more available (chest computerised tomography scanner – CT-scan) are highly sensitive (showing highly suggestive or quasi-pathognomonic lesions) for COVID-19 [29]. Practitioners therefore started to use chest CT-scan as (etiologic) diagnostic test for COVID-19. That tendency was so important that the Minister of Health (MoH) had to warn against abusive and inappropriate use of chest CT-scan as diagnostic tests for COVID-19 [29, 32].

For 73.6% of respondents, biologic test was compulsory to confirm COVID-19 and the figure rose to 96.4% among physicians (Table 2). That position is in line with current international practice guidelines for diagnosis of COVID-19 [33]. The long delays between admission of suspect cases in the COVID-19 isolation units and the results of RT-PCR have led to the numerous conflicts between family members and health staff at the BRH [30, 34, 35]. In contrast with rich countries where HCWs have been praised for their commitment against COVID-19, they have been assaulted in Bafoussam and elsewhere in Cameroon like in others LMICs under suspicion of (i) spreading the disease (ii) giving false positive diagnosis and (iii) preventing ritual funerals for COVID-19 deceased patients [11, 34]. In other to restore serenity between HCWs and users of health services, the Cameroonian Head of State had to take position in favour of previous in a public address to the nation [36].

Half (53.6%) of respondents (with no significant difference between categories) rightly answered that all currents treatment regimens for covid-19 were exclusively supportive [33]. This explains why only a third of them believed in the efficacy of chloroquine/hydroxychloroquine (CQ/HCQ), azithromycin and zinc that are the main molecules recommended by the MoH [37]. A striking finding is that one third (31.9%) of respondents thought that CQ/HCQ is effective in preventing COVID-19. This undoubtedly contributed to the massive off-the-counter use of (standard and sub-standard) CQ/HCQ leading to stock-outs of in pharmacies and illegal production reported few weeks after the outbreak of the epidemic in Cameroon. Several authors have warned against potential misuse and lack of medicines against covid-19 in Africa [7, 29, 38]. Furthermore a third (36.9%) of respondents thought that (anti-malarial) herbal remedies are protective against COVID-19. They certainly contributed to their widespread prophylactic and therapeutic consumption despite several warnings by health authorities [29]. Indeed, the alleged efficacy CQ/HCQ against COVID-19 and its recommendation by health authorities and scientific bodies in many rich countries led many people including opinion leaders to deduct that other anti-malarial drugs (artemisinin and amino-quinoleins derivatives) are also effective [39, 40].

If more than half of HCWs in a referral hospital are convinced that modern (so-called *white man medicine)* medicine has no curative solution for a novel pandemic like COVID-19 then it is understandable that most of lay Cameroonians look for solutions in their ancestral traditional pharmacopeia that is used for 80% health needs in SSA and in Cameroon in particular [41]. Half of HCWs didn’t know if HAART (Highly active antiretroviral therapy for People living with HIV/AIDS-PLWHA) has a protective effect against severe forms of COVID-19 and only 12.2% thought it was protective. Given that most of respondents (74.7%) knew that COVID-19 is not specific to elder and frail individuals, one can safely deduct than they will not wrongly reassure PLWHA that they are protected against COVID-19 by their routine HAART. To date there is no evidence of efficacy of antiretrovirals against COVID-19 [29, 42].

Management of COVID-19 corpses in Cameroon has frequently degenerates into violent conflicts between health authorities and families mainly because the latter are neither given the corpses nor allowed to carry out ritual funerals (toileting of the corpses, autopsies, funerals gatherings and religious ceremonies) [35]. HCWs are front liners in implementing this governmental measure because corpses are kept in mortuaries before burial by others public administrations (councils, police and gendarmerie). If all categories of HCWs of the BRH agreed that all COVID-19 related deaths occurring out of health facilities must immediately be reported to authorities and that COVID-19 corpses should be buried within the shortest possible delay, they differed on the roles and pre-eminence of authorities and families. For 87.5% of respondents, authorities should take an active part in handling COVID-19 corpses but only 44.1% of respondents thought that the dignity and cultural and religious traditions of the deceased and their family must be respected throughout. WHO guidance on the issue are clearly against hasty disposal of corpses from COVID-19 and stand in favour of the rights of the families as far post-mortem infection prevention and control measures are observed [43]. Recently research is being focused on autopsies in post mortem COVID- 19 cases. In the BRH a unit is currently researching on the “COVID-19 lung”. Ethical issues in this domain which are traditional in this community are accentuated in the case of COVID-19 for obvious reasons.

Almost all respondents (96.3%) acknowledged the severity of COVID-19; this is very important for their involvement in the facility-based response to the pandemic [12]. More than three quarters (78.8%) of respondents felt they were sufficiently compliant with HBDM measures while on duty but the significant difference (p = 0.02) observed indicates that the figures were higher for administrative and WASH personnel than for clinicians and biologists who work closer to patients. This is understandable given that (contrary to the previous) the latter are routinely in close contact with patients and their relatives in a context where communities don’t respect HBDM as perceived by 79.9% of respondents (Table 3). Nurses in China also reported excessive exposure to SARS-CoV-2 due to contact with patients [19]. Poor compliance to HBDM in communities in Bafoussam and elsewhere in Cameroon has been repeatedly condemned by high level officials (including the Head of Government) [44]. Abdelaziz et al. also reported that HCWs depicted poor adhesion to HBDM by population in central Maghreb (Tunisia, Morocco and Algeria) [12].

In Africa HCWs have influential voices in their communities and societies and are powerful agents of change, this is why their behaviours during epidemics are considered by lay people as indicators of the reality and severity of the condition [6]. With respect to that, HCWs of the BRH behave in an exemplary manner as 85.5% of them said they strictly apply HBDM in daily life and 93.7% of them discuss COVID-19 issues with their communities. This is another reason why they should be properly trained (theoretically and practically) and motivated to ensure community ownership of HBDM and successful local actions against COVID-19 [7, 12].

Though three quarters of health staff are either concerned or worried about the pandemic a close proportion (71.1%) is optimistic about the future of the pandemic in their region. That optimism of HCWs is quite important for communities given the context marked by repeated alarming and alarmist COVID-19 related predictions on media. In a qualitative survey, HCWs in Central Maghreb attributed the success of COVID-19 response strategies to the remarkable involvement of health professionals and communities [12].

More than three quarters of respondents reported a significant drop in their routine practice. In several other settings routine health services have been interrupted because hospitals were overwhelmed by COVID-19 surge [45]. The BRH was not overwhelmed by COVID-19 patients but populations out of fear of nosocomial contamination tended to avoid and/or postpone non urgent hospital visits or opt for alternative (often sub-standard) solutions. This led to a worrying drop in essential services for non-COVID-19 diseases. Other causes of disruptions of primary health care services in emerging epidemics are the disruption and transfers of resources. Several ruling bodies have warned against the drama of losing hard-won gains in controlling infections and non-infectious diseases in SSA in face of the COVID-19 pandemonium [1, 2, 6, 7, 45].

It was striking to observe that more than half of frontline HCWs (55.6%) were not satisfied with the response of their hospital to the covid-19 pandemic (Table 3). Their main grievances (Figure 1) pertained to the following categories: medicines and technologies (74.6%), service delivery (28.1%) and human resource (10.9%). Moreover 70% of staffs said they were not knowledgeable enough on how to manage COVID-19 patients, while only 32% of them felt that their protection (PPE) on duty was good. Indeed, good attitudes towards HBDM alone cannot ensure adequate protection of HCW which depend upon availability of personal protective equipment (hydro-alcoholic gel, facial mask, gear, goggle, gloves, boots, etc…) that was lacking. Huang et al. reported insufficient knowledge and skills among nurses in a Chinese COVID-19 hospital and Semaan et al. found that only 47% of HCWs from LMICs reported adequate training to manage COVID-19 cases [19, 45].

For 85.6% of staffs the BRH didn’t have the required resources (human, material, financial, institutional) for an optimal response to COVID-19. This reflects unpreparedness and slow responsiveness of the BRH and the Cameroonian health system to a surge of COVID-19 cases. Lack of personal protective equipment, medicines (CQ/HCQ were removed from the local market two decades ago), laboratory tests, ventilators and lack of regular updated guidelines for practices have been reported in LMICs and in SSA [1, 2, 7, 45]. Though high levels of infection of HCWs in charge of COVID-19 patients have been reported worldwide, shortages of personal protective equipment and laboratory tests have been pointed as causative of the disproportionate infection of HCWs in SSA with respect to the share of the continent in the global burden of the pandemic [45]. At the time of writing, systematic testing of HCWs was not done at the BRH to assess the prevalence of COVID-19. Indeed despite vigorous and voluntarist decisions implemented at continental level under the leadership of African Union and its Centre for Diseases

Control at the onset of the pandemic, health systems in the majority of SSA countries were too weak to adequately respond to the pandemic [1, 2, 4, 6, 7,12].

Though the global scientific community currently agrees that a vaccine will be the only long term solution to COVID-19, more than half of staffs of the BRH were against the idea of hosting a COVID-19 vaccine trial in their Region [47, 48]. This reveals deeply rooted misconceptions about immunization and clinical research. Indeed, their motivations (Figure 2) are either groundless (conspiracy theory (22.4%), irrelevance of a COVID-19 vaccine (10.7%) and public health threat by vaccine trials(33.7%) or not directed against vaccine trials themselves (mistrust in the government to properly monitor such sensitive clinical research (33.2%)). In case of need, those reasons can easily be overcome by a skilful communication campaign and good governance [49, 50].

This study is the first to provide an insight in knowledge and perception of HCW regarding COVID-19 in West-Cameroon and was carried out among HCWs in the unique COVID-19 treatment hospital of the region. It can therefore serve as an indirect “internal” audit of the hospital-based response to the pandemic in Cameroon. The single-hospital nature of this study limits the extrapolation of the results. Another major limit of this study lies in the content and the structure of the questionnaire that was drafted by authors thus resulting in a certain degree of subjectivity of the survey results; nonetheless is worth noting that reliable, valid, standardized and context-specific questionnaires for COVID-19 are not yet available.

## Conclusions

Knowledge of COVID-19 among Health Care Workers of the Bafoussam Regional Hospital was good regarding clinical signs and biological diagnosis but was poor regarding transmission, therapeutics and management of corpses. Their perception of the hospital response to the pandemic was unfavorable depicting several shortcomings to address: lack of medicines and technologies, inadequate service delivery and shortage of skilled human resources. A significant drop of non-COVID-19 services has been reported by respondents. Findings suggest more resources must be allocated to improve the facility-based response to COVID-19 at the Bafoussam Regional Hospital. Given that the COVID-19 pandemic is here to stay, knowledge of HCWs must be routinely updated and practice guidelines should be made available to them. Special attention must be paid to the continuum of essential services for non-COVID-19 diseases. Current Cameroonian regulation on management of COVID-19 corpses should be made more culture-sensitive. The great popularity of herbal remedies (from Cameroonian pharmacopeia) for COVID-19 among HCWs deserves special attention by health authorities.

## Data Availability

All data supporting this manuscript are available on request.

## Acknowledgments

Authors are grateful to all the staff of the Bafoussam Regional Hospital for their participation. They also thank Tamukum Moses for his valuable technical support.

## Notes

### Competing Interest Statement

The authors have declared no competing interest.

### Funding Statement

no external funding was received

### Author Declarations

The Bafoussam Regional Hospital Institutional review board has approved this study

